# Weighted burden analysis in 200,000 exome-sequenced subjects characterises rare variant effects on risk of type 2 diabetes

**DOI:** 10.1101/2021.01.08.21249453

**Authors:** David Curtis

## Abstract

Type 2 diabetes (T2D) is a disease for which both common genetic variants and environmental factors influence risk. A few genes have been identified in which very rare variants have large effects on risk and here we carry out a weighted burden analysis of rare variants in a sample of over 200,000 exome-sequenced participants in the UK Biobank project, of whom over 13,000 have T2D. Variant weights were allocated based on allele frequency and predicted effect, as informed by a previous analysis of hyperlipidaemia. There was an exome-wide significant increased burden of rare, functional variants in three genes, *GCK, HNF4A* and *GIGYF1. GIGYF1* has not previously been identified as a diabetes risk gene but its product is plausibly involved in the modification of insulin signalling. A number of other genes did not attain exome-wide significance but were highly ranked and potentially of interest, including *ALAD, PPARG, GYG1* and *GHRL*. Loss of function (LOF) variants were associated with T2D in *GCK* and *GIGYF1* whereas nonsynonymous variants annotated as probably damaging were associated in *GCK* and *HNF4A*. Overall, fewer than 1% of T2D cases carried one of these variants. In two genes previously implicated in diabetes aetiology, *HNF1A* and *HNF1B*, there was an excess of LOF variants among cases but the small numbers of these fell well short of statistical significance, suggesting that even larger datasets will be helpful for more fully elucidating the contribution of rare genetic variants to T2D risk. This research has been conducted using the UK Biobank Resource.

## Introduction

Genome-wide association studies of type 2 diabetes (T2D) have implicated a large number of common genetic variants (Xue et al., 2018). In the UK Biobank, a genetic risk score derived from common variants was associated with T2D and incorporating it alongside conventional risk factors in order to predict T2D increased the area under the curve (AUC) from 0.851 to 0.855 (Chen et al., 2021). Common variants have individually modest effects on risk but a small number of genes have been identified in which rare coding variants with large effects can result in a phenotype of non-insulin dependent diabetes. Maturity onset diabetes of the young (MODY) is caused by mutations in a number of genes including *HNF1A, HNF4A, GCK, HNF1B, KCNJ11, ABCC8* (Murphy et al., 2008). Loss of function (LOF) and nonsynonymous variants in *PPARG* can cause a familial lipodystrophy with insulin resistant diabetes (Agostini et al., 2018). Recessively acting mutations in the *INSR* gene can cause insulin resistance with hyperglycaemia (Semple et al., 2011). These rare coding variants have typically been identified by targeted studies of familial cases and individuals with severe phenotypes but the availability of large samples of exome sequenced subjects now allows the possibility to explore the effects of variations in these genes in the population more broadly and potentially to discover novel genes. A study of 16,000 exome and genome sequenced cases and controls failed to detect a substantial contribution from rare variants to risk of type 2 diabetes (Fuchsberger et al., 2016). However a larger study using 20,791 cases and 24,440 controls identified three exome-wide significant genes, *MC4R, SLC30A8* and *PAM*, as well as specific variant in *MC4R*, rs79783591 (Ile269Asn), which was exome-wide significant in single variant analyses (Flannick et al., 2019).

Exome sequence data is now available for 200,000 of the 500,000 UK Biobank subjects (Szustakowski et al., 2020). We have recently analysed this in order to illuminate the effect of rare, coding variants on susceptibility to hyperlipidaemia and we now apply the same approach to study T2D (Curtis, 2021a).

## Methods

The UK Biobank dataset was downloaded along with the variant call files for 200,632 subjects who had undergone exome-sequencing and genotyping by the UK Biobank Exome Sequencing Consortium using the GRCh38 assembly with coverage 20X at 95.6% of sites on average (Szustakowski et al., 2020). UK Biobank had obtained ethics approval from the North West Multi-centre Research Ethics Committee which covers the UK (approval number: 11/NW/0382) and had obtained informed consent from all participants. The UK Biobank approved an application for use of the data (ID 51119) and ethics approval for the analyses was obtained from the UCL Research Ethics Committee (11527/001). All variants were annotated using the standard software packages VEP, PolyPhen and SIFT (Adzhubei et al., 2013; Kumar et al., 2009; McLaren et al., 2016). To obtain population principal components reflecting ancestry, version 2.0 of *plink* (https://www.cog-genomics.org/plink/2.0/) was run with the options *--maf 0*.*1 --pca 20 approx* (Chang et al., 2015; Galinsky et al., 2016).

The UK Biobank sample contains 503,317 subjects of whom 94.6% are of white ethnicity. As we have discussed previously, it has become standard practice for investigators to simply discard data from participants with other ancestries and we regard this as regrettable (Curtis, 2021b). We demonstrated that if population principal components are included as covariates then it is possible to include all participants, regardless of ancestry, in the type of weighted burden analysis described here without any inflation of the test statistic.

To define cases, a similar approach was used as was previously implemented for the investigation of hyperlipidaemia (Curtis, 2021a, 2019). The T2D phenotype was determined from three sources in the dataset: self-reported diabetes or type 2 diabetes (but not type 1 or gestational diabetes); reporting taking any of a list of named medications commonly used to treat T2D in the UK (https://www.diabetes.co.uk/Diabetes-drugs.html); having an ICD10 code for non-insulin-dependent diabetes mellitus in hospital records or as a cause of death. Subjects in any of these categories were deemed to be cases while all other subjects were taken to be controls.

The SCOREASSOC program was used to carry out a weighted burden analysis to test whether, in each gene, sequence variants which were rarer and/or predicted to have more severe functional effects occurred more commonly in cases than controls. Attention was restricted to rare variants with minor allele frequency (MAF) <= 0.01 in both cases and controls. As previously described, variants were weighted by overall MAF so that variants with MAF=0.01 were given a weight of 1 while very rare variants with MAF close to zero were given a weight of 10 (Curtis, 2021b). Variants were also weighted according to their functional annotation using the GENEVARASSOC program, which was used to generate input files for weighted burden analysis by SCOREASSOC (Curtis, 2016, 2012). The weights were informed from the analysis of the effects of different categories of variant in *LDLR* on hyperlipidaemia risk (Curtis, 2021a). Variants predicted to cause complete loss of function (LOF) of the gene were assigned a weight of 100. Nonsynonymous variants were assigned a weight of 5 but if PolyPhen annotated them as possibly or probably damaging then 5 or 10 was added to this and if SIFT annotated them as deleterious then 20 was added. In order to allow exploration of the effects of different types of variant on disease risk the variants were also grouped into broader categories to be used in multivariate analyses as described below. The full set of weights and categories is displayed in Table 1. As described previously, the weight due to MAF and the weight due to functional annotation were multiplied together to provide an overall weight for each variant. Variants were excluded if there were more than 10% of genotypes missing in the controls or if the heterozygote count was smaller than both homozygote counts in the controls. If a subject was not genotyped for a variant then they were assigned the subject-wise average score for that variant. For each subject a gene-wise weighted burden score was derived as the sum of the variant-wise weights, each multiplied by the number of alleles of the variant which the given subject possessed. For variants on the X chromosome, hemizygous males were treated as homozygotes.

**Table 1.** The table shows the weight which was assigned to each type of variant as annotated by VEP, Polyphen and SIFT as well as the broad categories which were used for multivariate analyses of variant effects (Adzhubei et al., 2013; Kumar et al., 2009; McLaren et al., 2016).

| **VEP / SIFT / Polyphen annotation** | **Weight** | **Category** |
| --- | --- | --- |
| intergenic_variant | 0 | Unused |
| feature_truncation | 0 | Intronic, etc. |
| regulatory_region_variant | 0 | Intronic, etc. |
| feature_elongation | 0 | Intronic, etc. |
| regulatory_region_amplification | 1 | Intronic, etc. |
| regulatory_region_ablation | 1 | Intronic, etc. |
| TF_binding_site_variant | 1 | Intronic, etc. |
| TFBS_amplification | 1 | Intronic, etc. |
| TFBS_ablation | 1 | Intronic, etc. |
| downstream_gene_variant | 0 | Intronic, etc. |
| upstream_gene_variant | 0 | Intronic, etc. |
| non_coding_transcript_variant | 0 | Intronic, etc. |
| NMD_transcript_variant | 0 | Intronic, etc. |
| intron_variant | 0 | Intronic, etc. |
| non_coding_transcript_exon_variant | 0 | Intronic, etc. |
| 3_prime_UTR_variant | 1 | 3 prime UTR |
| 5_prime_UTR_variant | 1 | 5 prime UTR |
| mature_miRNA_variant | 5 | Unused |
| coding_sequence_variant | 0 | Unused |
| synonymous_variant | 0 | Synonymous |
| stop_retained_variant | 5 | Unused |
| incomplete_terminal_codon_variant | 5 | Unused |
| splice_region_variant | 1 | Splice region |
| protein_altering_variant | 5 | Protein altering |
| missense_variant | 5 | Protein altering |
| inframe_deletion | 10 | InDel, etc |
| inframe_insertion | 10 | InDel, etc |
| transcript_amplification | 10 | InDel, etc |
| start_lost | 10 | Unused |
| stop_lost | 10 | Unused |
| frameshift_variant | 100 | Disruptive |
| stop_gained | 100 | Disruptive |
| splice_donor_variant | 100 | Splice site variant |
| splice_acceptor_variant | 100 | Splice site variant |
| transcript_ablation | 100 | Disruptive |
| SIFT deleterious | 20 | Deleterious |
| PolyPhen possibly damaging | 5 | Possibly damaging |
| PolyPhen probably damaging | 10 | Probably damaging |

For each gene, logistic regression analysis was carried out including the first 20 population principal components and sex as covariates and a likelihood ratio test was performed comparing the likelihoods of the models with and without the gene-wise burden score. The statistical significance was summarised as a signed log p value (SLP), which is the log base 10 of the p value given a positive sign if the score is higher in cases and negative if it is higher in controls.

Gene set analyses were carried out as before using the 1454 “all GO gene sets, gene symbols” pathways as listed in the file *c5*.*all*.*v5*.*0*.*symbols*.*gmt* downloaded from the Molecular Signatures Database at http://www.broadinstitute.org/gsea/msigdb/collections.jsp (Subramanian et al., 2005). For each set of genes, the natural logs of the gene-wise p values were summed according to Fisher’s method to produce a chi-squared statistic with degrees of freedom equal to twice the number of genes in the set. The p value associated with this chi-squared statistic was expressed as a minus log10 p (MLP) as a test of association of the set with the hyperlipidaemia phenotype.

For selected genes, additional analyses were carried out to clarify the contribution of different categories of variant. As described previously, logistic regression analyses were performed on the counts of the separate categories of variant as listed in Table 1, again including principal components and sex as covariates, to estimate the effect size for each category (Curtis, 2021a). The odds ratios associated with each category were estimated along with their standard errors and the Wald statistic was used to obtain a p value, except for categories in which variants occurred fewer than 50 times in which case Fisher’s exact test was applied to the variant counts. The associated p value was converted to an SLP, again with the sign being positive if the mean count was higher in cases than controls.

Data manipulation and statistical analyses were performed using GENEVARASSOC, SCOREASSOC and R (R Core Team, 2014).

## Results

There were 13,938 cases of T2D and 186,694 controls. There were 20,384 genes for which there were qualifying variants. Given that there were 20,384 informative genes, the critical threshold for the absolute value of the SLP to declare a result as formally statistically significant is -log10(0.05/20384) = 5.61 and this was achieved by three genes, *GCK* (SLP = 22.24), *HNF4A* (SLP = 6.82) and *GIGYF1* (SLP = 6.22). The quantile-quantile (QQ) plot for the SLPs obtained for all genes except *GCK* is shown in Figure 1. This shows that the test appears to be well-behaved and conforms well with the expected distribution. Omitting the genes with the 100 highest and 100 lowest SLPs, which might be capturing a real biological effect, the gradient for positive SLPs is 1.06 with intercept at -0.007 and the gradient for negative SLPs is 1.02 with intercept at -0.009, indicating only modest inflation of the test statistic in spite of the fact that participants of all ancestries are included.

**Figure 1.**
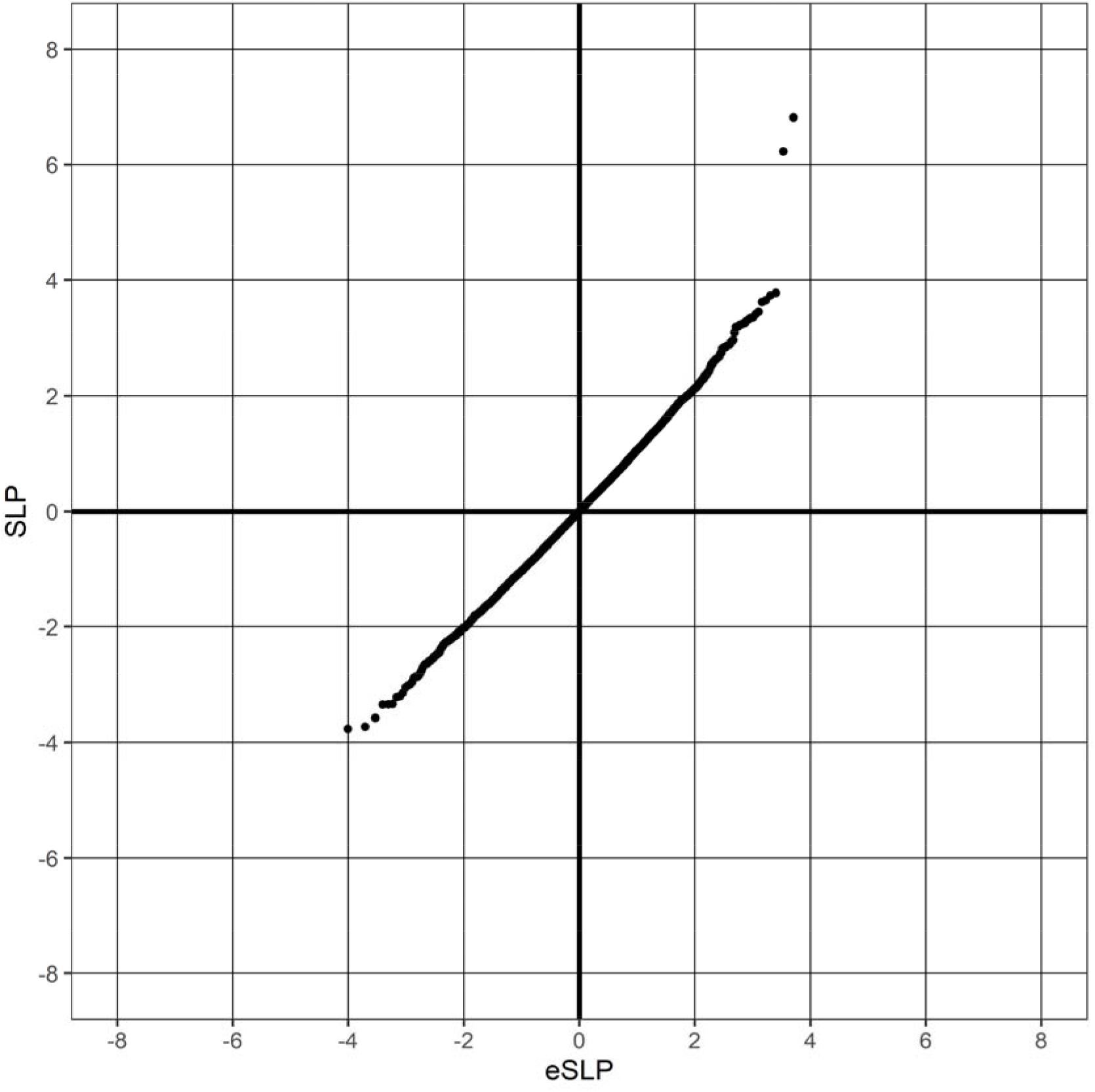
QQ plot of SLPs obtained for weighted burden analysis of association with hyperlipidaemia showing observed against expected SLP for each gene, omitting results for GCK, which has SLP = 22.25.

Variants in *GCK* are recognised as the cause of up to half of cases of MODY, itself accounting for around 1-2% of cases of all diabetes diagnoses (Bishay and Greenfield, 2016). Likewise, *HNF4A* variants cause 5-10% of cases of MODY (Naylor et al., 2018). By contrast, *GIGYF1* has not previously been implicated in the aetiology of diabetes although it is known that its product binds to growth factor receptor-bound protein 10 (GRB10) and has a role in modulating the insulin-like growth factor (IGF1) receptor signalling pathway (Giovannone et al., 2003; Zhou et al., 2018). A variant in *GRB10* has been reported to be associated with decreased early-phase insulin secretion and the muscle-specific ablation of *Grb10* in mice causes increased glucose uptake into muscles with increased insulin signalling (Holt et al., 2018; Lyssenko et al., 2015). However *GRB10* itself showed no evidence for association in T2D in the current analysis (SLP = -0.02).

Table 2 shows all the genes achieving SLP with absolute value greater than 3, equivalent to an uncorrected p value of 0.001. Given that 20,384 genes were tested, one would expect that by chance about 20 would reach this level of significance whereas in fact there are 32. Thus it is possible that some of these highly ranked genes do demonstrate a biological signal which fails to reach statistical significance after correction for multiple testing and some of them seem worth commenting on. The expression of *ALAD* (SLP = 3.63) is reduced in obese subjects while the expression of Alad is reduced in rats with high-fat diet-induced weight gain (Moreno-Navarrete et al., 2017). Additionally, inhibition of ALAD with aminotriazole led to reduced glucose uptake in cultured human adipocytes. The common P12A variant of *PPARG* (SLP = 3.45) reduces risk of T2D whereas rare LOF variants and nonsynonymous variants which cause reduced activity (occurring in approximately 1 in 1,000 individuals) substantially increase risk (Majithia et al., 2014). Damaging variants in *GYG1* (SLP = 3.22) cause deficiency of glycogenin 1, resulting in glycogen storage myopathies, but have not been reported to be associated with diabetes (Ben Yaou et al., 2017). *GHRL* (SLP = -3.15) encodes the ghrelin-obestatin preproprotein which is cleaved to yield two peptides, ghrelin and obestatin, which are involved in appetite and energy metabolism and there have been some studies which have claimed that the common Leu72Met (rs696217) variant is associated with reduced risk of T2D although the effect does not seem to be consistent and the gene was not highlighted in a large GWAS meta-analysis (Rivera-León et al., 2020; Xue et al., 2018). The results for all sets are provided in Supplementary Table S1.

**Table 2.** Genes with absolute value of SLP exceeding 3 or more (equivalent to p<0.001) for test of association of weighted burden score with T2D.

| Symbol | SLP | Name |
| --- | --- | --- |
| GCK | 22.25 | Glucokinase |
| HNF4A | 6.82 | Hepatocyte Nuclear Factor 4 Alpha |
| GIGYF1 | 6.22 | GRB10 Interacting GYF Protein 1 |
| ZNF620 | 3.78 | Zinc Finger Protein 620 |
| RAI2 | 3.74 | Retinoic Acid Induced 2 |
| TM4SF20 | 3.65 | Transmembrane 4 L Six Family Member 20 |
| ALAD | 3.63 | Aminolevulinate Dehydratase |
| PPARG | 3.45 | Peroxisome Proliferator Activated Receptor Gamma |
| LOC105370752 | 3.42 | Uncharacterized LOC105370752 |
| KLHL11 | 3.35 | Kelch Like Family Member 11 |
| HMGXB4 | 3.35 | HMG-Box Containing 4 |
| MIR6825 | 3.31 | MicroRNA 6825 |
| TAZ | 3.30 | Tafazzin |
| WDR33 | 3.25 | WD Repeat Domain 33 |
| HECTD1 | 3.24 | HECT Domain E3 Ubiquitin Protein Ligase 1 |
| ZNF571-AS1 | 3.23 | ZNF571 Antisense RNA 1 |
| GYG1 | 3.22 | Glycogenin 1 |
| APTX | 3.20 | Aprataxin |
| KCNK15 | 3.19 | Potassium Two Pore Domain Channel Subfamily K Member 15 |
| XPO1 | 3.19 | Exportin 1 |
| PKD1 | 3.10 | Polycystin 1, Transient Receptor Potential Channel Interacting |
| ZNF763 | -3.01 | Zinc Finger Protein 763 |
| COA5 | -3.05 | Cytochrome C Oxidase Assembly Factor 5 |
| GHRL | -3.15 | Ghrelin And Obestatin Prepropeptide |
| DEUP1 | -3.20 | Deuterosome Assembly Protein 1 |
| C7orf50 | -3.22 | Chromosome 7 Open Reading Frame 50 |
| MFSD12 | -3.34 | Major Facilitator Superfamily Domain Containing 12 |
| C19orf73 | -3.34 | Chromosome 19 Open Reading Frame 73 |
| ATXN1L | -3.35 | Ataxin 1 Like |
| EML4 | -3.58 | EMAP Like 4 |
| DLEC1 | -3.72 | DLEC1 Cilia And Flagella Associated Protein |
| RPS5 | -3.76 | Ribosomal Protein S5 |

In order to see if any additional genes were highlighted by analysing gene sets, gene set analysis was performed as described above after first removing all genes with absolute SLP value greater than 3. Given that 1,454 sets were tested a critical MLP to achieve to declare results significant after correction for multiple testing would be log10(1454*20) = 4.46 and this was not achieved by any set. There were two sets with MLP > 3, EXTERNAL SIDE OF PLASMA MEMBRANE (MLP = 3.29) and MONOSACCHARIDE TRANSMEMBRANE TRANSPORTER ACTIVITY (MLP = 3.06). The latter is of some interest because it consists of 10 genes, of which three were individually significant at p<0.05, these being *SLC2A2* (SLP = 2.54), *SLC2A3* (SLP = -2.37) and *SLC2A4* (SLP = 1.70). *SLC2A2*, previously known as *GLUT2*, codes for a glucose transporter expressed by beta cells which senses glucose levels and recessively acting variants in it can cause neonatal diabetes (Sansbury et al., 2012). A common intronic variant of *SLC2A2*, rs8192675, is associated with the glycaemic response to metformin (Zhou et al., 2016). *SLC2A4* codes for a glucose transporter whose levels in cell membranes increase in response to insulin but although candidate gene studies claim that common variants in it are associated with T2D these results are not supported by properly powered GWAS metanalysis (Hu et al., 2019; Xue et al., 2018). The results for all sets are provided in Supplementary Table S2.

For the genes of possible interest listed above, a logistic regression analysis of different categories of variant was carried out to elucidate their relative contributions. The results for the three exome-wide significant genes are shown in Table 3, which shows differences between the genes relating to the implicated pattern of variants. The results for *GCK* demonstrate that splice site variants and gene disruptive variants, comprising frameshift and stop variants, are associated with large effects on risk. These occur a total of 17 times among the 13,938 cases. However of note is that nonsynonymous variants annotated as probably damaging by PolyPhen are also associated with increased risk, with OR = 2.97 (1.59 - 5.54), and these occur 33 times among cases. The situation for *HNF4A* is quite different. There are no splice site variants and only 6 gene disruptive variants and these all occur in controls. Only probably damaging nonsynonymous variants show an effect, with OR = 2.97 (1.61 - 5.50), and these occur 34 times among cases. Finally, for *GIGYF1* probably damaging nonsynonymous variants have no discernible effect and it is only the splice site (OR = 7.70 (2.62 - 22.67)) and disruptive (OR = 5.65 (3.07 - 10.40)) variants which increase risk and these occur 24 times in cases.

**Table 3.**
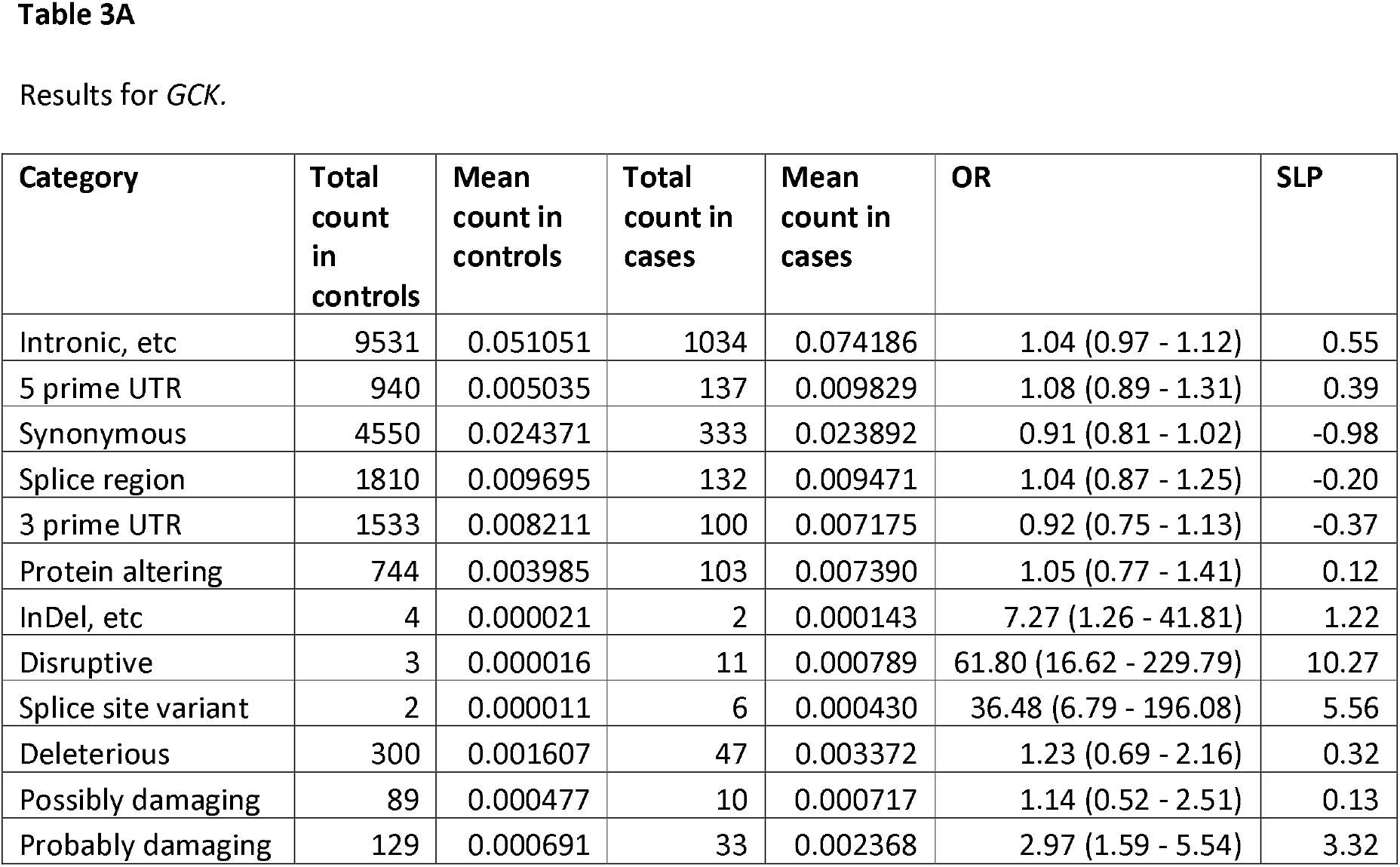
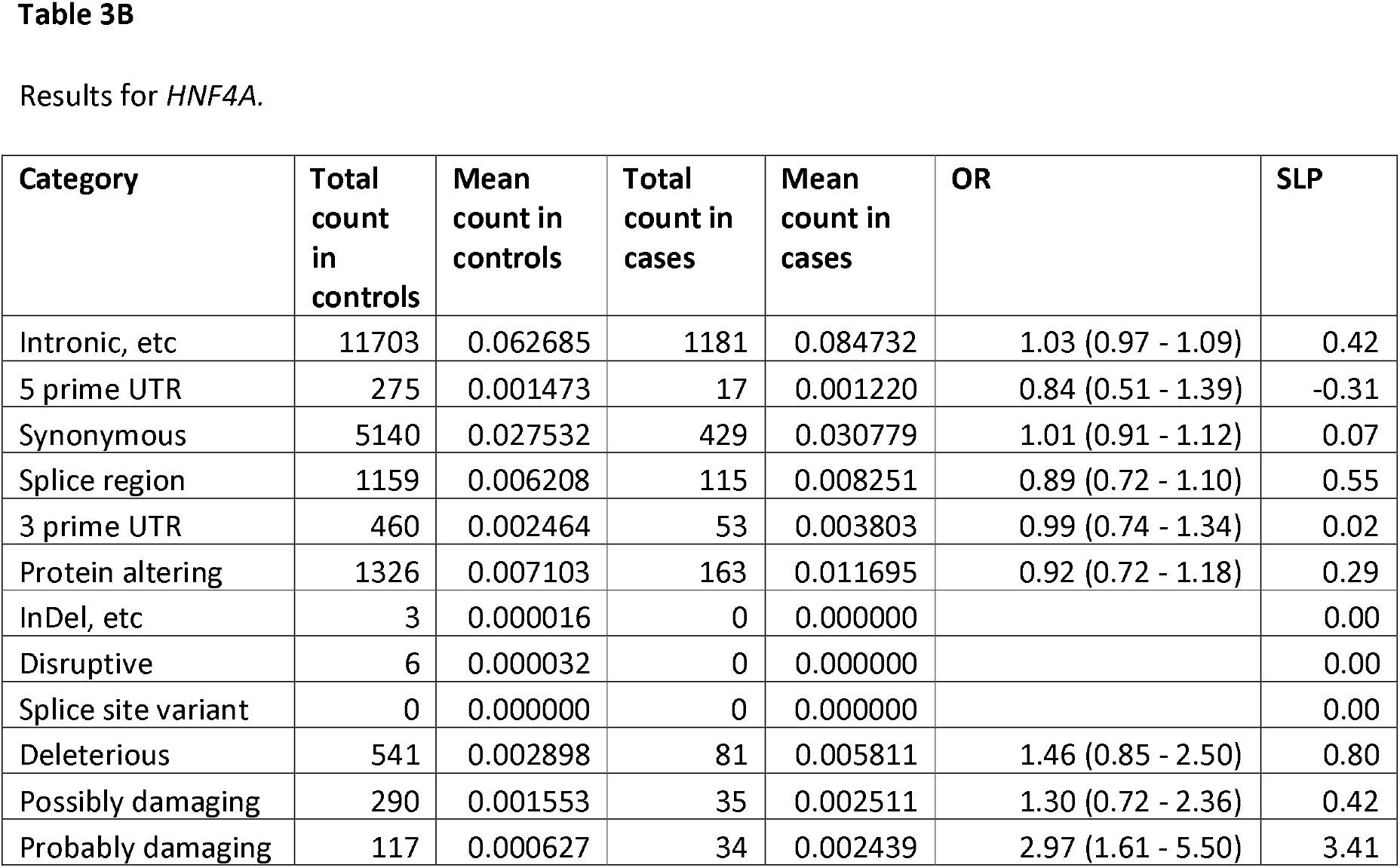
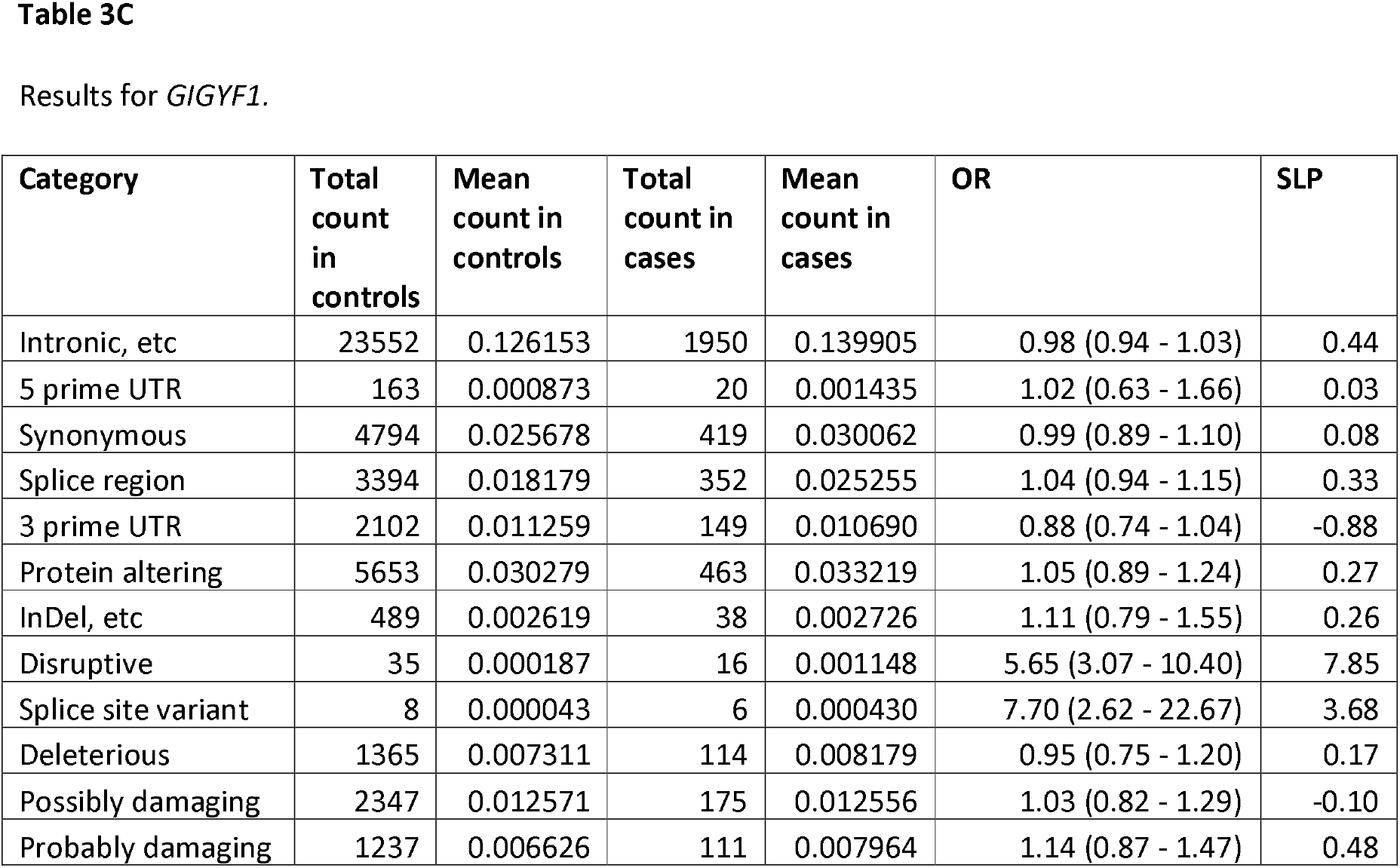
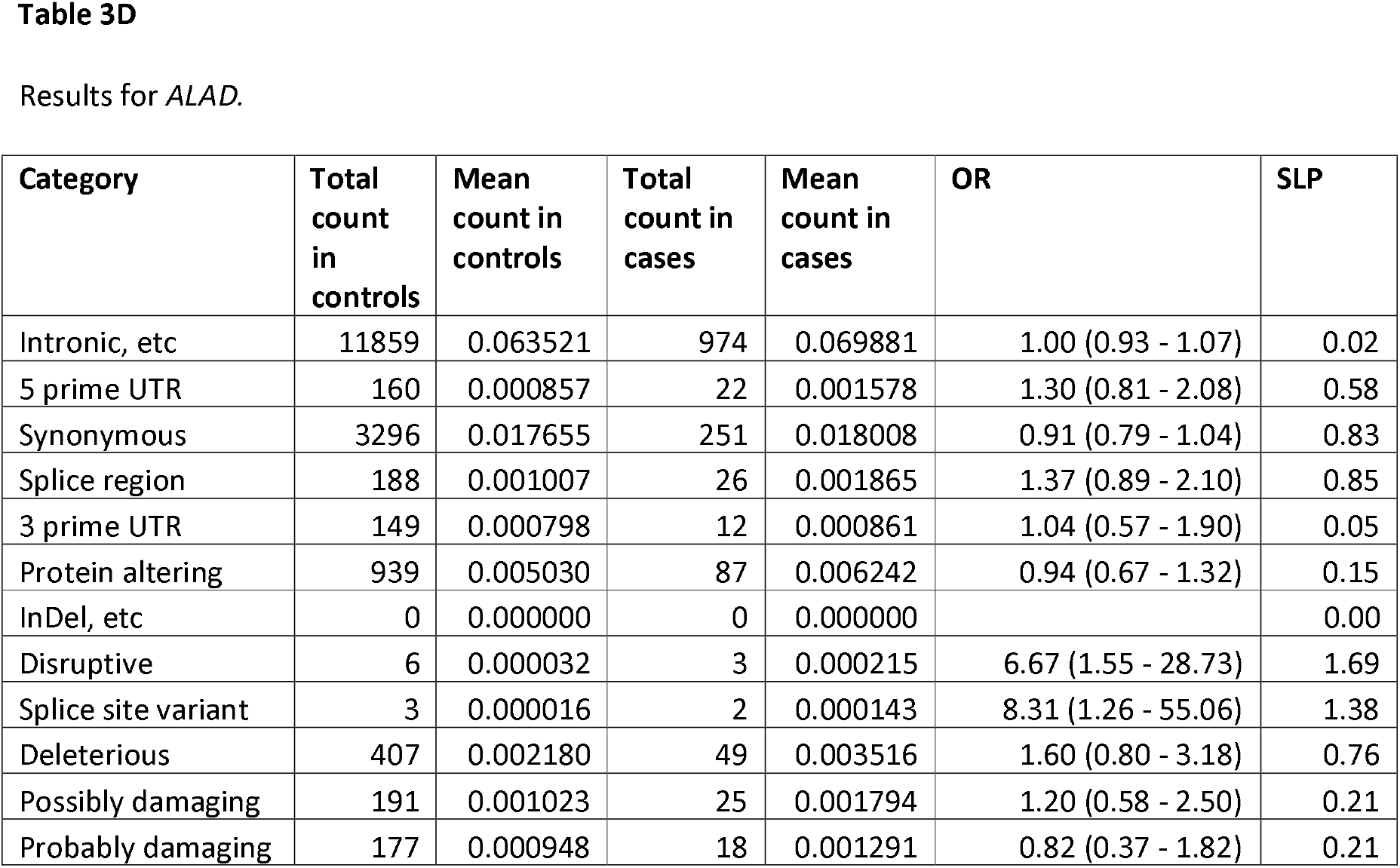
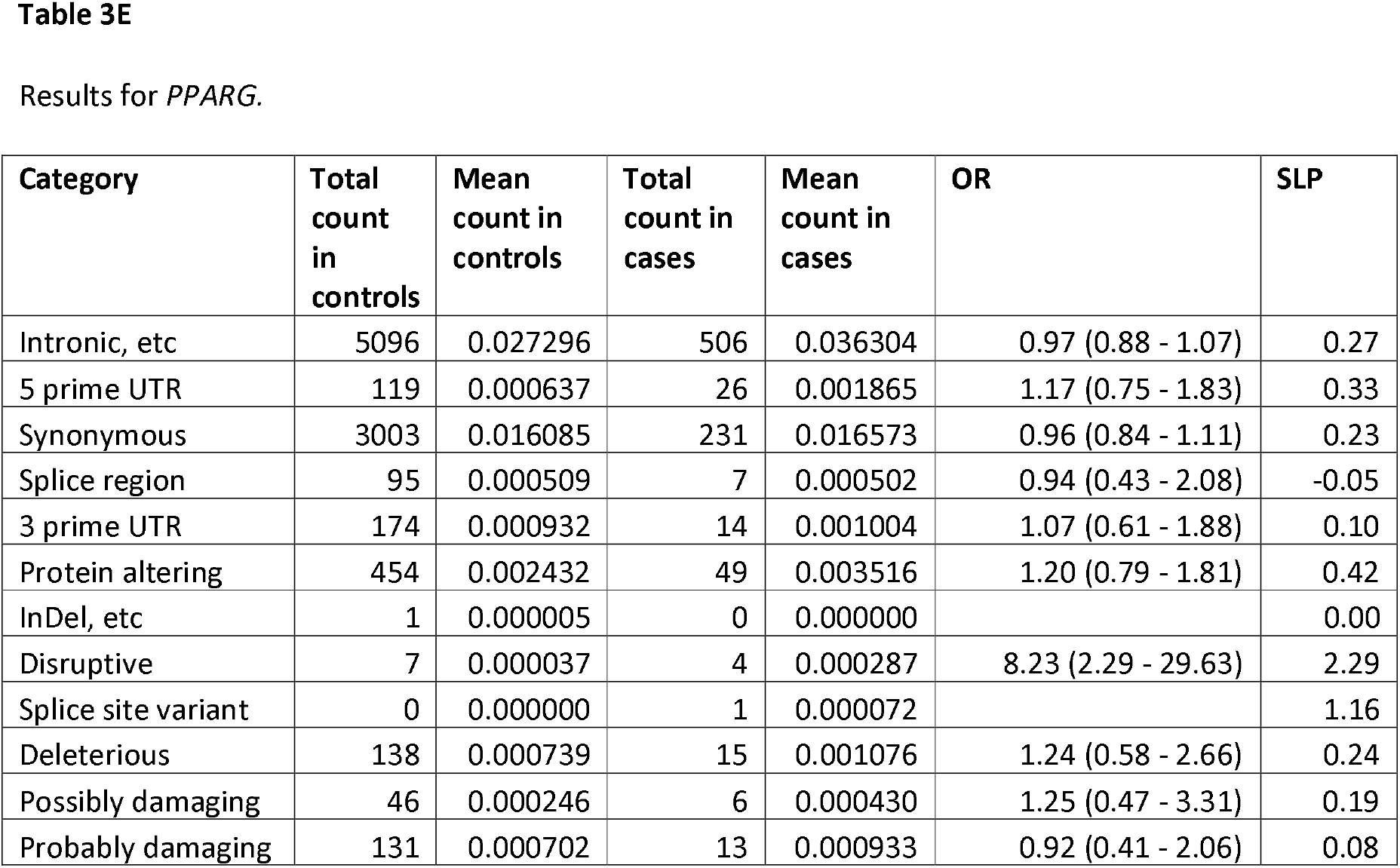
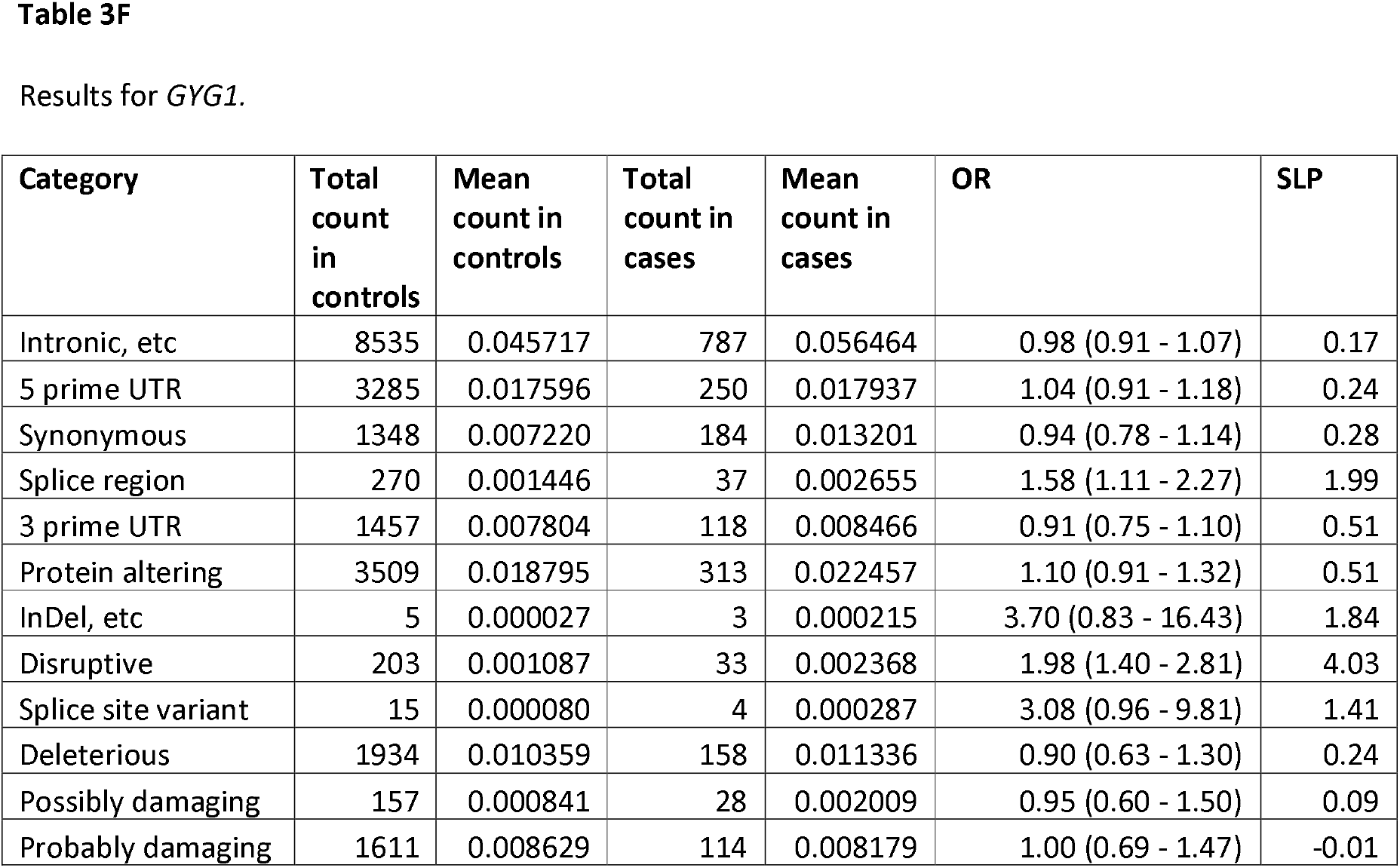
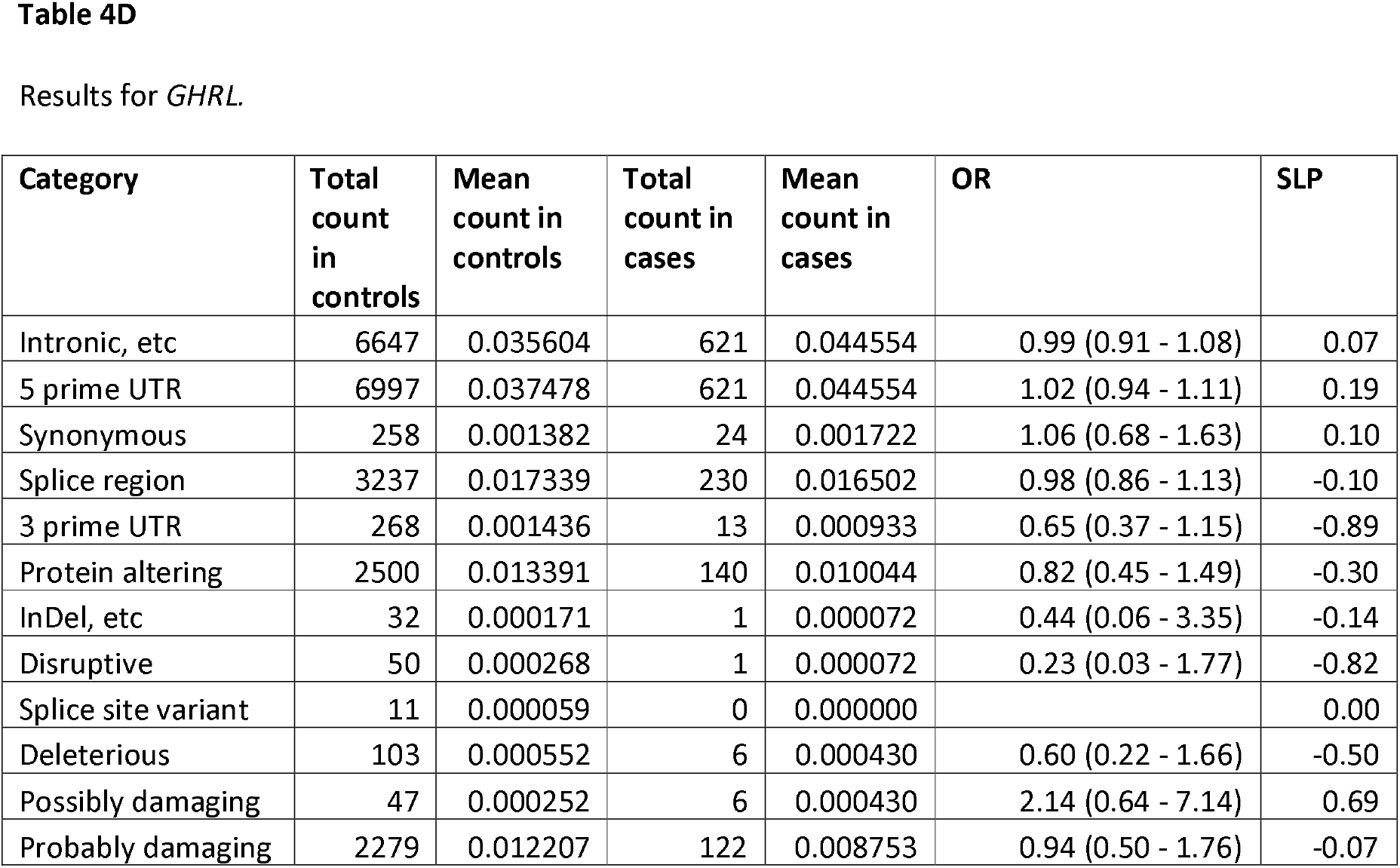
Results from logistic regression analysis showing the effects on risk of T2D of different categories of variant within exome-wide significant genes and genes of interest with absolute value of gene-wise SLP > 3. Odds ratios for each category are estimated including principal components and sex as covariates.

| Category | Total count in controls | Mean count in controls | Total count in cases | Mean count in cases | OR | SLP |
| --- | --- | --- | --- | --- | --- | --- |
| Intronic, etc | 9531 | 0.051051 | 1034 | 0.074186 | 1.04 (0.97 - 1.12) | 0.55 |
| 5 prime UTR | 940 | 0.005035 | 137 | 0.009829 | 1.08 (0.89 - 1.31) | 0.39 |
| Synonymous | 4550 | 0.024371 | 333 | 0.023892 | 0.91 (0.81 - 1.02) | -0.98 |
| Splice region | 1810 | 0.009695 | 132 | 0.009471 | 1.04 (0.87 - 1.25) | -0.20 |
| 3 prime UTR | 1533 | 0.008211 | 100 | 0.007175 | 0.92 (0.75 - 1.13) | -0.37 |
| Protein altering | 744 | 0.003985 | 103 | 0.007390 | 1.05 (0.77 - 1.41) | 0.12 |
| InDel, etc | 4 | 0.000021 | 2 | 0.000143 | 7.27 (1.26 - 41.81) | 1.22 |
| Disruptive | 3 | 0.000016 | 11 | 0.000789 | 61.80 (16.62 - 229.79) | 10.27 |
| Splice site variant | 2 | 0.000011 | 6 | 0.000430 | 36.48 (6.79 - 196.08) | 5.56 |
| Deleterious | 300 | 0.001607 | 47 | 0.003372 | 1.23 (0.69 - 2.16) | 0.32 |
| Possibly damaging | 89 | 0.000477 | 10 | 0.000717 | 1.14 (0.52 - 2.51) | 0.13 |
| Probably damaging | 129 | 0.000691 | 33 | 0.002368 | 2.97 (1.59 - 5.54) | 3.32 |
| Intronic, etc | 11703 | 0.062685 | 1181 | 0.084732 | 1.03 (0.97 - 1.09) | 0.42 |
| 5 prime UTR | 275 | 0.001473 | 17 | 0.001220 | 0.84 (0.51 - 1.39) | -0.31 |
| Synonymous | 5140 | 0.027532 | 429 | 0.030779 | 1.01 (0.91 - 1.12) | 0.07 |
| Splice region | 1159 | 0.006208 | 115 | 0.008251 | 0.89 (0.72 - 1.10) | 0.55 |
| 3 prime UTR | 460 | 0.002464 | 53 | 0.003803 | 0.99 (0.74 - 1.34) | 0.02 |
| Protein altering | 1326 | 0.007103 | 163 | 0.011695 | 0.92 (0.72 - 1.18) | 0.29 |
| InDel, etc | 3 | 0.000016 | 0 | 0.000000 |  | 0.00 |
| Disruptive | 6 | 0.000032 | 0 | 0.000000 |  | 0.00 |
| Splice site variant | 0 | 0.000000 | 0 | 0.000000 |  | 0.00 |
| Deleterious | 541 | 0.002898 | 81 | 0.005811 | 1.46 (0.85 - 2.50) | 0.80 |
| Possibly damaging | 290 | 0.001553 | 35 | 0.002511 | 1.30 (0.72 - 2.36) | 0.42 |
| Probably damaging | 117 | 0.000627 | 34 | 0.002439 | 2.97 (1.61 - 5.50) | 3.41 |
| Intronic, etc | 23552 | 0.126153 | 1950 | 0.139905 | 0.98 (0.94 - 1.03) | 0.44 |
| 5 prime UTR | 163 | 0.000873 | 20 | 0.001435 | 1.02 (0.63 - 1.66) | 0.03 |
| Synonymous | 4794 | 0.025678 | 419 | 0.030062 | 0.99 (0.89 - 1.10) | 0.08 |
| Splice region | 3394 | 0.018179 | 352 | 0.025255 | 1.04 (0.94 - 1.15) | 0.33 |
| 3 prime UTR | 2102 | 0.011259 | 149 | 0.010690 | 0.88 (0.74 - 1.04) | -0.88 |
| Protein altering | 5653 | 0.030279 | 463 | 0.033219 | 1.05 (0.89 - 1.24) | 0.27 |
| InDel, etc | 489 | 0.002619 | 38 | 0.002726 | 1.11 (0.79 - 1.55) | 0.26 |
| Disruptive | 35 | 0.000187 | 16 | 0.001148 | 5.65 (3.07 - 10.40) | 7.85 |
| Splice site variant | 8 | 0.000043 | 6 | 0.000430 | 7.70 (2.62 - 22.67) | 3.68 |
| Deleterious | 1365 | 0.007311 | 114 | 0.008179 | 0.95 (0.75 - 1.20) | 0.17 |
| Possibly damaging | 2347 | 0.012571 | 175 | 0.012556 | 1.03 (0.82 - 1.29) | -0.10 |
| Probably damaging | 1237 | 0.006626 | 111 | 0.007964 | 1.14 (0.87 - 1.47) | 0.48 |
| Intronic, etc | 11859 | 0.063521 | 974 | 0.069881 | 1.00 (0.93 - 1.07) | 0.02 |
| 5 prime UTR | 160 | 0.000857 | 22 | 0.001578 | 1.30 (0.81 - 2.08) | 0.58 |
| Synonymous | 3296 | 0.017655 | 251 | 0.018008 | 0.91 (0.79 - 1.04) | 0.83 |
| Splice region | 188 | 0.001007 | 26 | 0.001865 | 1.37 (0.89 - 2.10) | 0.85 |
| 3 prime UTR | 149 | 0.000798 | 12 | 0.000861 | 1.04 (0.57 - 1.90) | 0.05 |
| Protein altering | 939 | 0.005030 | 87 | 0.006242 | 0.94 (0.67 - 1.32) | 0.15 |
| InDel, etc | 0 | 0.000000 | 0 | 0.000000 |  | 0.00 |
| Disruptive | 6 | 0.000032 | 3 | 0.000215 | 6.67 (1.55 - 28.73) | 1.69 |
| Splice site variant | 3 | 0.000016 | 2 | 0.000143 | 8.31 (1.26 - 55.06) | 1.38 |
| Deleterious | 407 | 0.002180 | 49 | 0.003516 | 1.60 (0.80 - 3.18) | 0.76 |
| Possibly damaging | 191 | 0.001023 | 25 | 0.001794 | 1.20 (0.58 - 2.50) | 0.21 |
| Probably damaging | 177 | 0.000948 | 18 | 0.001291 | 0.82 (0.37 - 1.82) | 0.21 |
| Intronic, etc | 5096 | 0.027296 | 506 | 0.036304 | 0.97 (0.88 - 1.07) | 0.27 |
| 5 prime UTR | 119 | 0.000637 | 26 | 0.001865 | 1.17 (0.75 - 1.83) | 0.33 |
| Synonymous | 3003 | 0.016085 | 231 | 0.016573 | 0.96 (0.84 - 1.11) | 0.23 |
| Splice region | 95 | 0.000509 | 7 | 0.000502 | 0.94 (0.43 - 2.08) | -0.05 |
| 3 prime UTR | 174 | 0.000932 | 14 | 0.001004 | 1.07 (0.61 - 1.88) | 0.10 |
| Protein altering | 454 | 0.002432 | 49 | 0.003516 | 1.20 (0.79 - 1.81) | 0.42 |
| InDel, etc | 1 | 0.000005 | 0 | 0.000000 |  | 0.00 |
| Disruptive | 7 | 0.000037 | 4 | 0.000287 | 8.23 (2.29 - 29.63) | 2.29 |
| Splice site variant | 0 | 0.000000 | 1 | 0.000072 |  | 1.16 |
| Deleterious | 138 | 0.000739 | 15 | 0.001076 | 1.24 (0.58 - 2.66) | 0.24 |
| Possibly damaging | 46 | 0.000246 | 6 | 0.000430 | 1.25 (0.47 - 3.31) | 0.19 |
| Probably damaging | 131 | 0.000702 | 13 | 0.000933 | 0.92 (0.41 - 2.06) | 0.08 |
| Intronic, etc | 8535 | 0.045717 | 787 | 0.056464 | 0.98 (0.91 - 1.07) | 0.17 |
| 5 prime UTR | 3285 | 0.017596 | 250 | 0.017937 | 1.04 (0.91 - 1.18) | 0.24 |
| Synonymous | 1348 | 0.007220 | 184 | 0.013201 | 0.94 (0.78 - 1.14) | 0.28 |
| Splice region | 270 | 0.001446 | 37 | 0.002655 | 1.58 (1.11 - 2.27) | 1.99 |
| 3 prime UTR | 1457 | 0.007804 | 118 | 0.008466 | 0.91 (0.75 - 1.10) | 0.51 |
| Protein altering | 3509 | 0.018795 | 313 | 0.022457 | 1.10 (0.91 - 1.32) | 0.51 |
| InDel, etc | 5 | 0.000027 | 3 | 0.000215 | 3.70 (0.83 - 16.43) | 1.84 |
| Disruptive | 203 | 0.001087 | 33 | 0.002368 | 1.98 (1.40 - 2.81) | 4.03 |
| Splice site variant | 15 | 0.000080 | 4 | 0.000287 | 3.08 (0.96 - 9.81) | 1.41 |
| Deleterious | 1934 | 0.010359 | 158 | 0.011336 | 0.90 (0.63 - 1.30) | 0.24 |
| Possibly damaging | 157 | 0.000841 | 28 | 0.002009 | 0.95 (0.60 - 1.50) | 0.09 |
| Probably damaging | 1611 | 0.008629 | 114 | 0.008179 | 1.00 (0.69 - 1.47) | -0.01 |
| Intronic, etc | 6647 | 0.035604 | 621 | 0.044554 | 0.99 (0.91 - 1.08) | 0.07 |
| 5 prime UTR | 6997 | 0.037478 | 621 | 0.044554 | 1.02 (0.94 - 1.11) | 0.19 |
| Synonymous | 258 | 0.001382 | 24 | 0.001722 | 1.06 (0.68 - 1.63) | 0.10 |
| Splice region | 3237 | 0.017339 | 230 | 0.016502 | 0.98 (0.86 - 1.13) | -0.10 |
| 3 prime UTR | 268 | 0.001436 | 13 | 0.000933 | 0.65 (0.37 - 1.15) | -0.89 |
| Protein altering | 2500 | 0.013391 | 140 | 0.010044 | 0.82 (0.45 - 1.49) | -0.30 |
| InDel, etc | 32 | 0.000171 | 1 | 0.000072 | 0.44 (0.06 - 3.35) | -0.14 |
| Disruptive | 50 | 0.000268 | 1 | 0.000072 | 0.23 (0.03 - 1.77) | -0.82 |
| Splice site variant | 11 | 0.000059 | 0 | 0.000000 |  | 0.00 |
| Deleterious | 103 | 0.000552 | 6 | 0.000430 | 0.60 (0.22 - 1.66) | -0.50 |
| Possibly damaging | 47 | 0.000252 | 6 | 0.000430 | 2.14 (0.64 - 7.14) | 0.69 |
| Probably damaging | 2279 | 0.012207 | 122 | 0.008753 | 0.94 (0.50 - 1.76) | -0.07 |

Table 3 also shows the results for the genes which, while not exome-wide significant, had SLP with magnitude >3 and which appeared to be potentially of interest, *ALAD, PPARG, GYG1* and *GHRL*. The signal for *ALAD* seems to be driven by the fact that although splice site and disruptive variants occur only a total of 5 times in cases this still makes them about 7 times as frequent as in controls. The signal for *PPARG* arises from the fact that disruptive variants occur 4 times in cases and 7 times in controls, OR = 8.23 (2.29 - 29.63). The findings for *GYG1* seem somewhat more robust, being based on an excess of both disruptive (OR = 1.98 (1.40 - 2.81)) and splice site (OR = 3.08 (0.96 - 9.81)) variants in cases, with a total of 37 occurrences. For *GHRL*, which has SLP = -3,15, implying that variants in it might be protective, no individual category has a statistically significant effect and there is more a general tendency for there to be fewer variants among cases which is spread over a number of categories, including 3 prime UTR, protein altering, indel, disruptive and deleterious.

The analyses described above failed to highlight a number of genes which have previously been implicated in T2D, comprising *HNF1A* (SLP = 1.66), *HNF1B* (SLP = -0.28), *ABCC8* (SLP = 1.94), *INSR* (SLP = -0.25), *MC4R* (SLP = 1.41), *SLC30A8* (SLP = -2.64) and *PAM* (SLP = 0.19). The association with T2D for each category of variant within these genes is shown in Table 4. From this it can be seen that LOF variants in *HNF1A* and *HNF1B* are commoner in cases than controls but that their absolute numbers are too small to produce statistically significant effects. By contrast, LOF variants have higher overall frequency in *ABCC8* and they have approximately equal frequencies in cases and controls. These results are consistent with reports that it is only activating variants in *ABCC8* result in diabetes whereas LOF variants can cause hyperinsulinaemia (De Franco et al., 2020). The overall frequencies of different category of nonsynonymous variant do not vary between cases and controls, possibly reflecting the inability of SIFT and PolyPhen to distinguish variants which have a gain of function effect. The frequency of LOF variants in *INSR* are similar in cases and controls, indicating that, although recessively acting variants can cause infantile hyperinsulinaemia followed by insulin dependent diabetes, the loss of function of a single copy of this gene has little discernible effect on risk of T2D (Semple et al., 2011). The results for *MC4R* suggest that variants which are disruptive or annotated as deleterious or damaging may be associated with a moderate increase in risk, with OR 1.2 – 1.5, but no individual category is statistically significant. Unfortunately genotypes for the single *MC4R* variant previously reported to be associated with T2D, rs79783591 producing an Ile269Asn substitution, were not provided in the supplied dataset even though it is clearly exonic and genotypes for nearby variants are available. The frequency of nonsynonymous variants annotated as deleterious in *SLC30A8* is higher in controls than cases, consistent with previous findings that missense variants in this gene typically result in reduced T2D risk, but again this is not statistically significant (Flannick et al., 2019). In *PAM* there is no real suggestion of any signal among the included variants. However the frequency criteria used excluded the two variants which had previously been individually implicated statistically and through functional studies (Thomsen et al., 2018). When these were analysed separately they both showed evidence for a small effect on risk, with rs35658696 (Asp563Gly) having SLP = 5.68, OR = 1.15 (1.08 – 1.21) and rs78408340 having SLP = 6.92, OR = 1.20 (1.13 – 1.28).

**Table 4.**
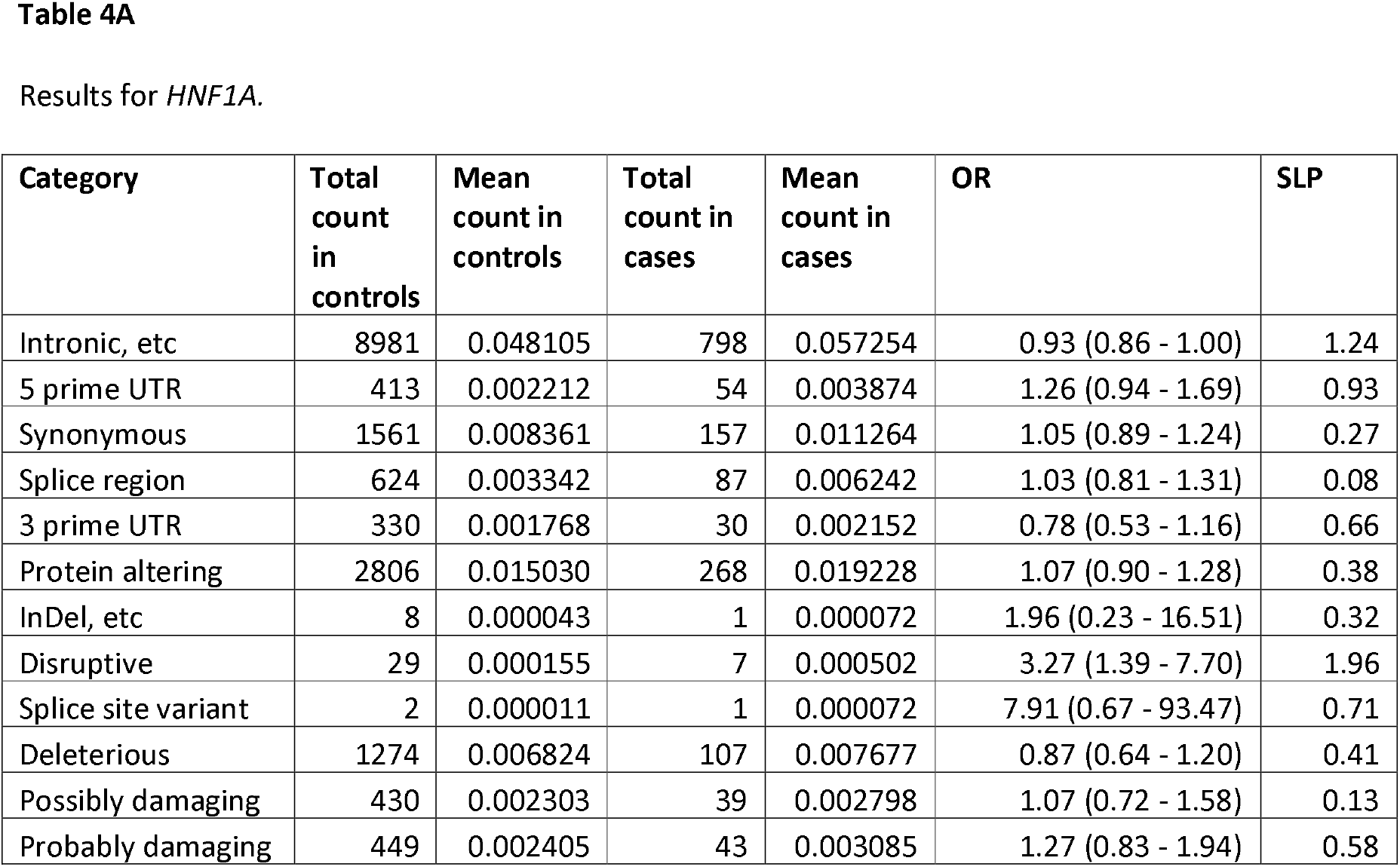
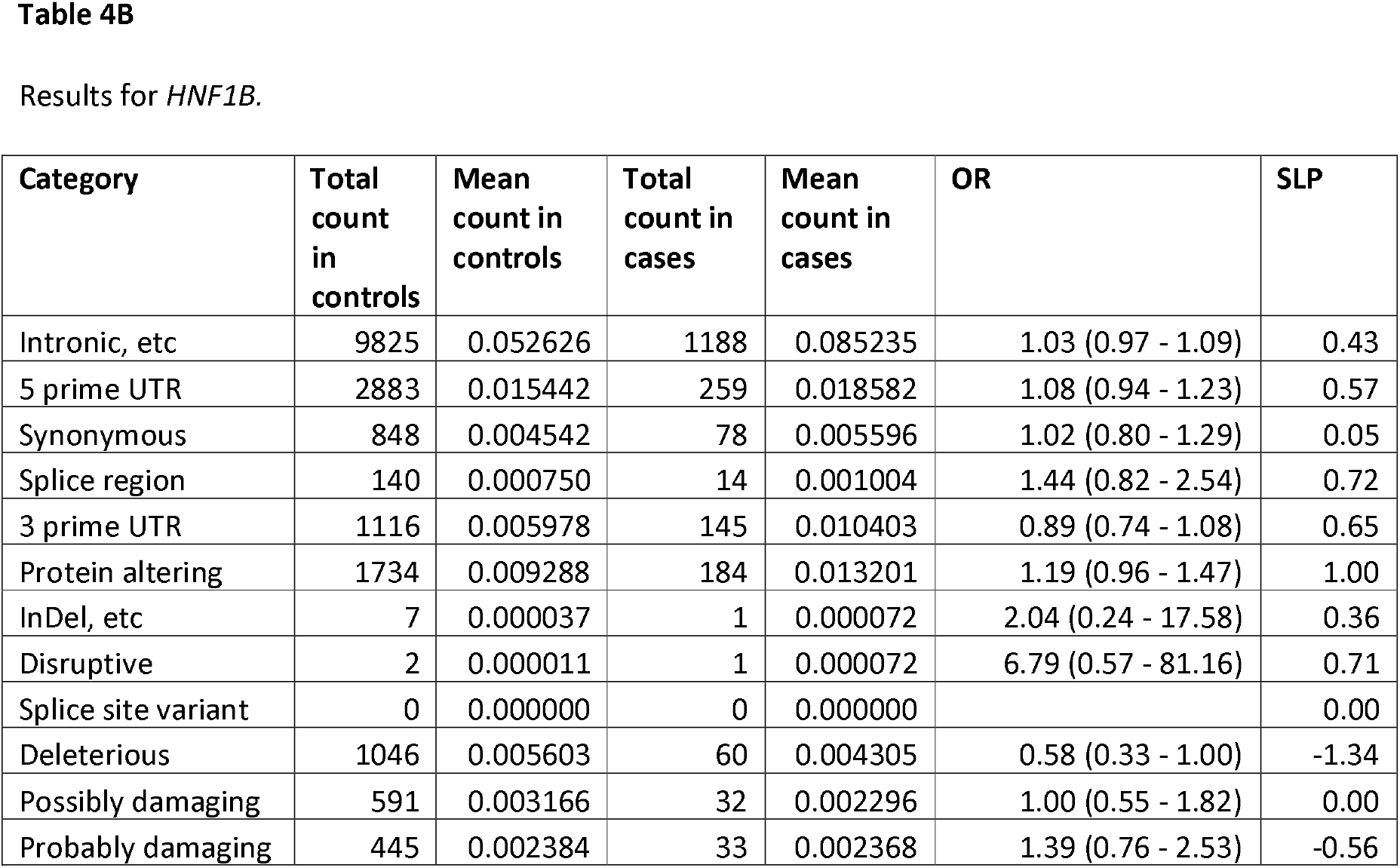
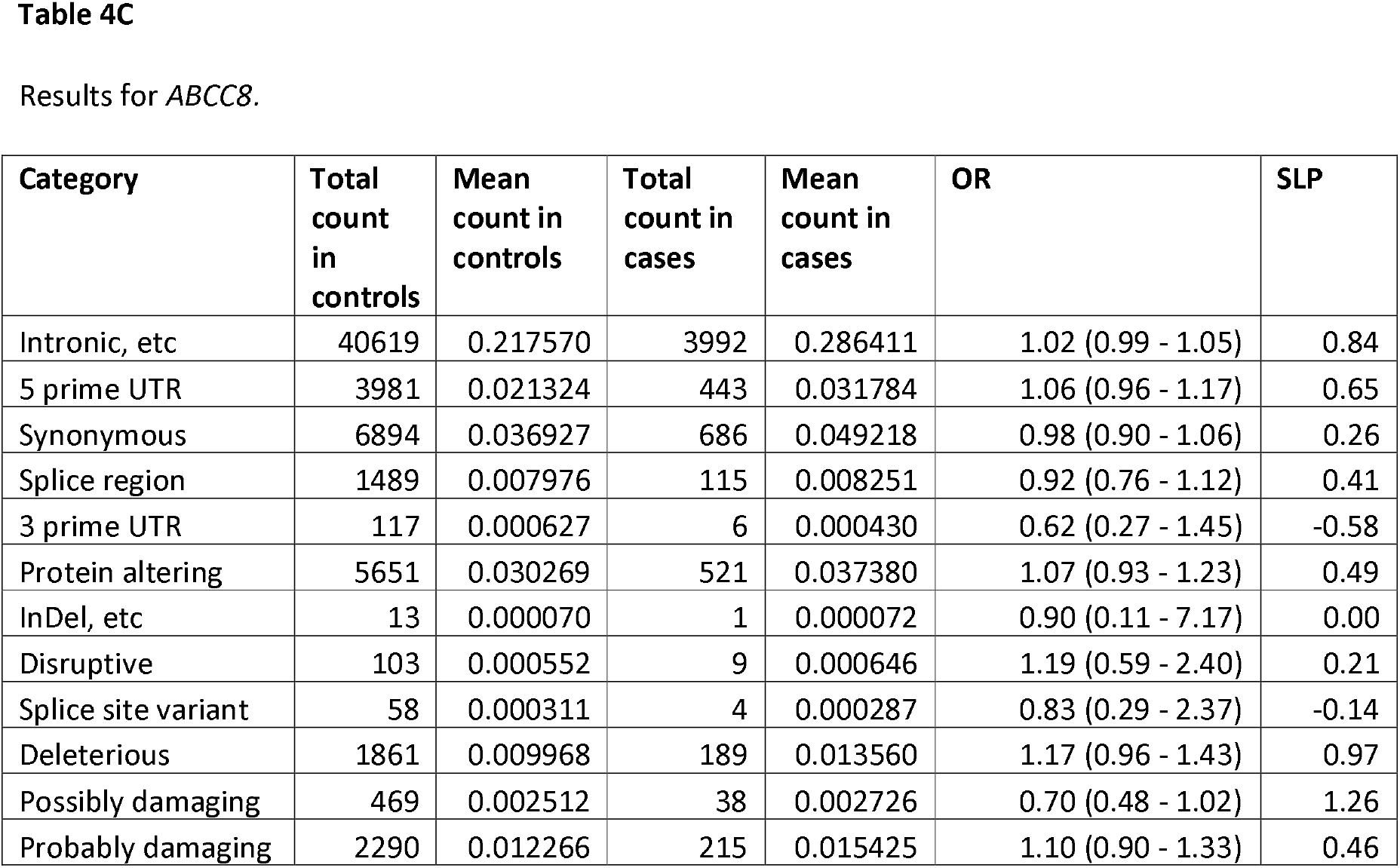
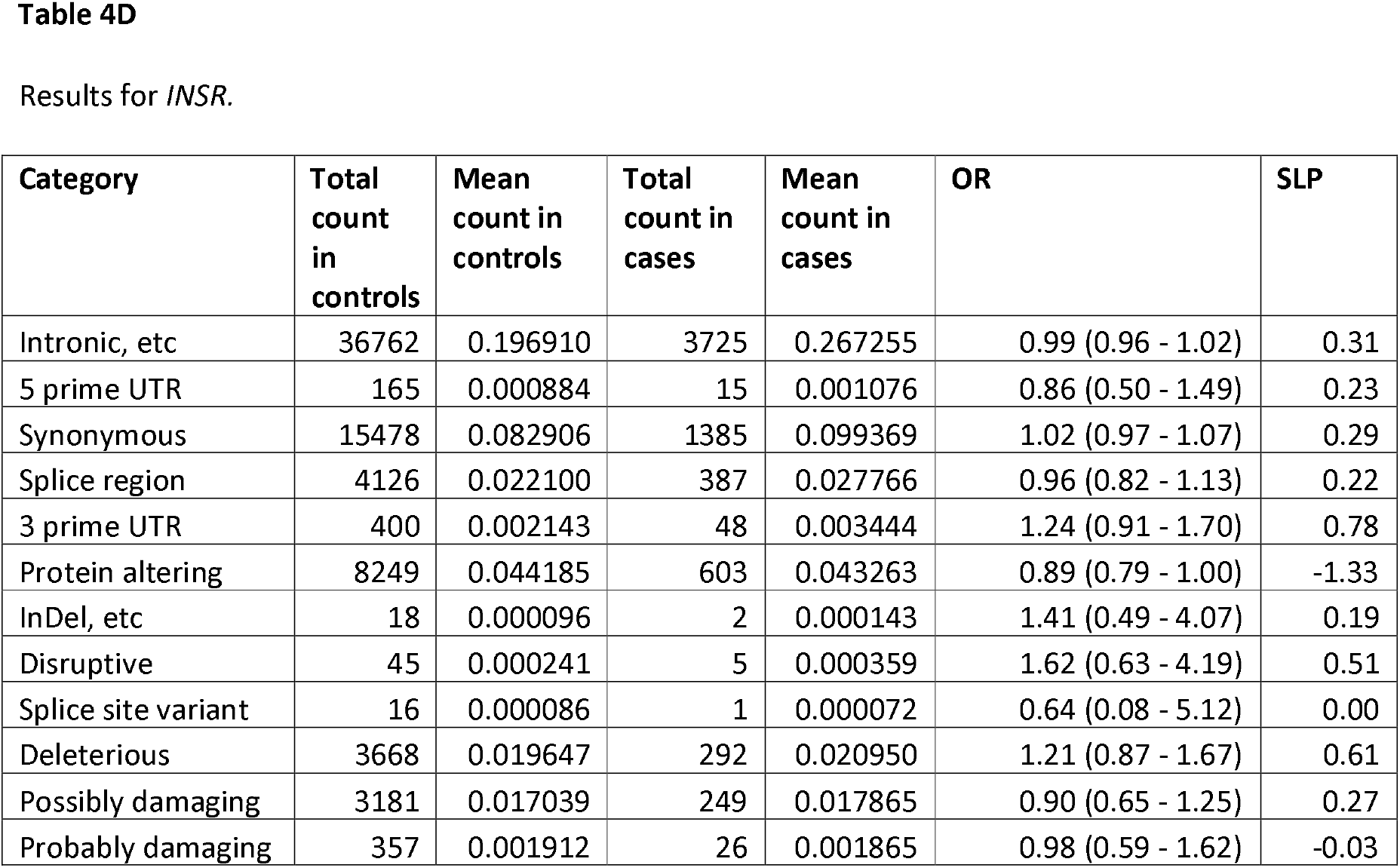
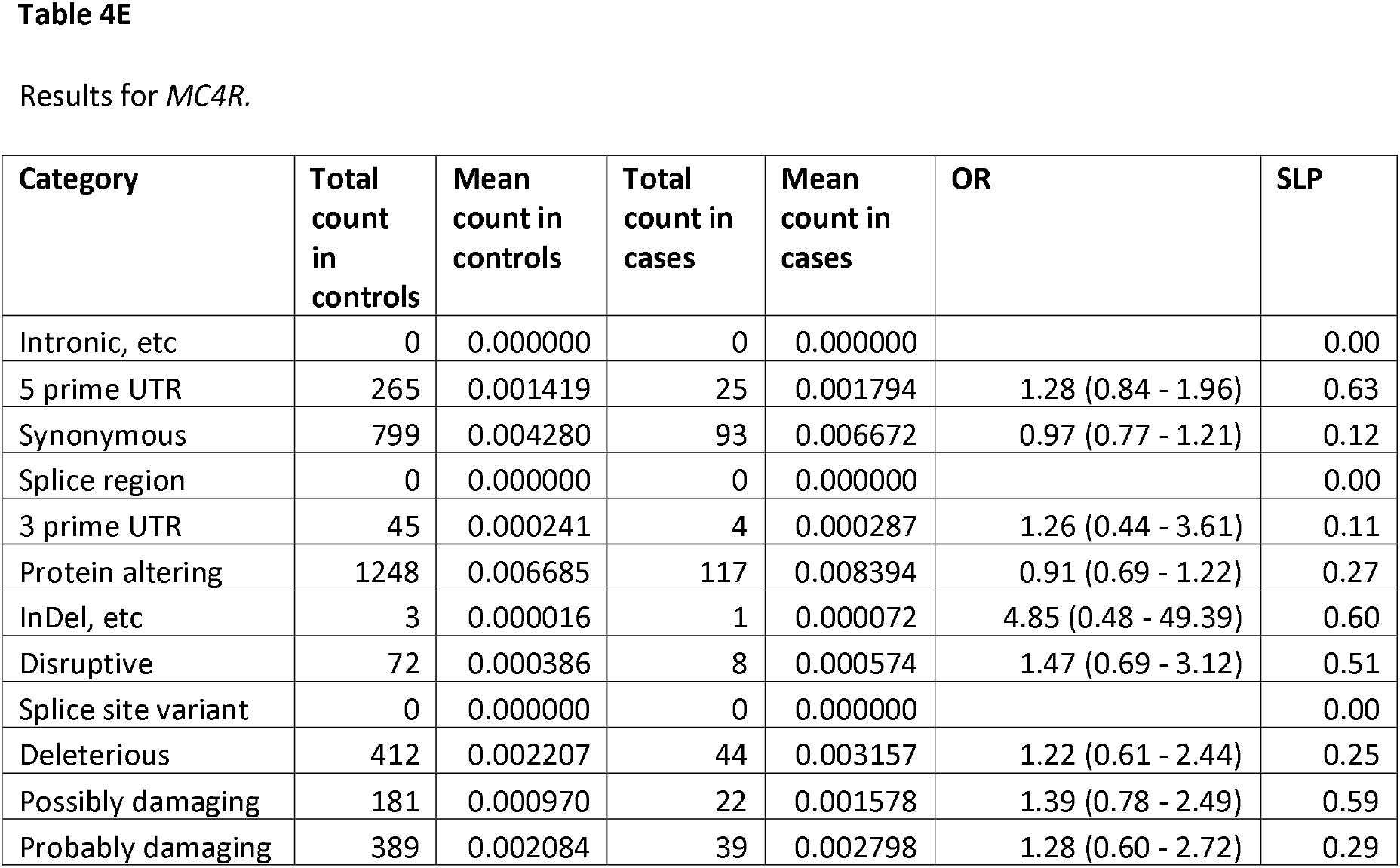
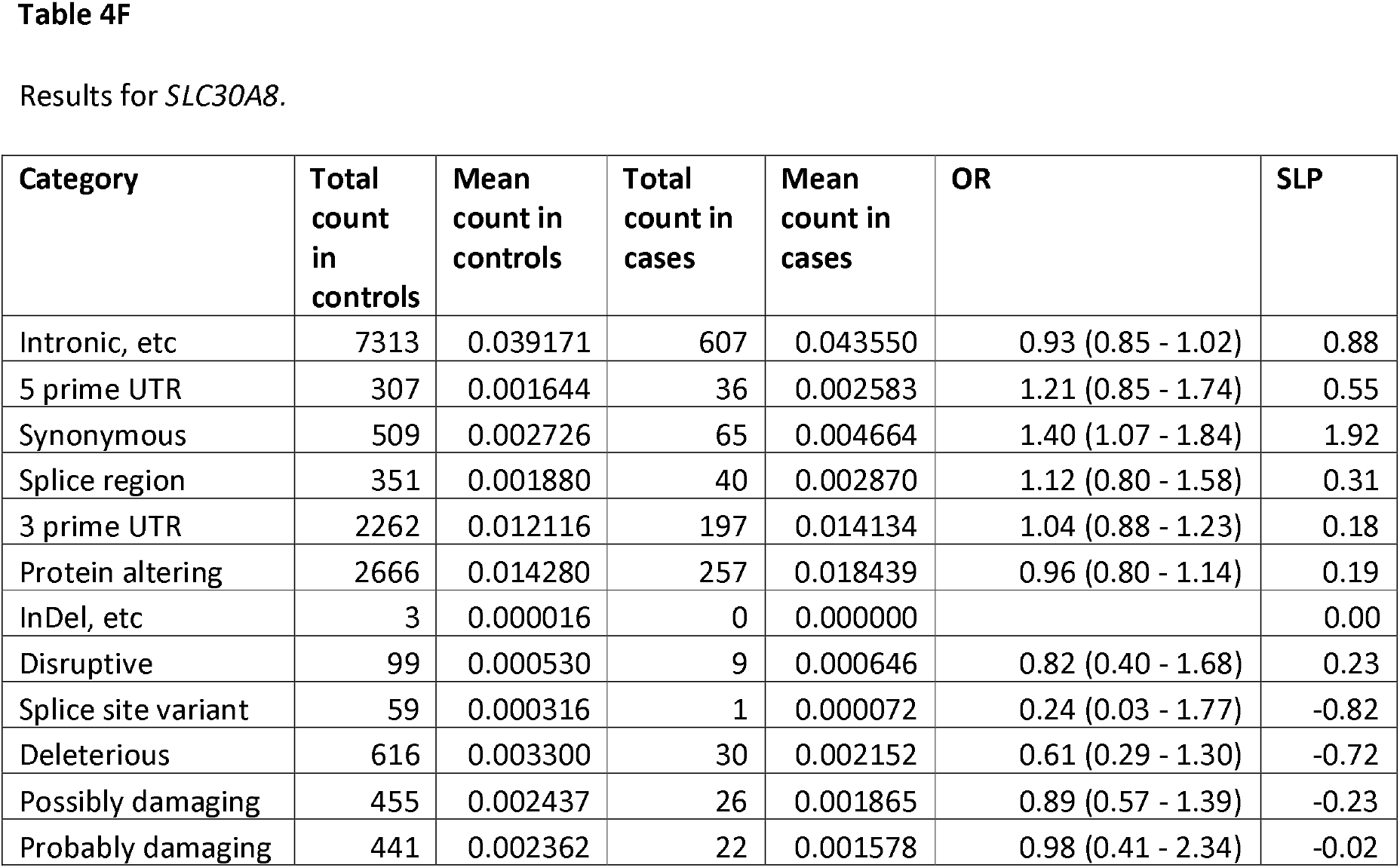
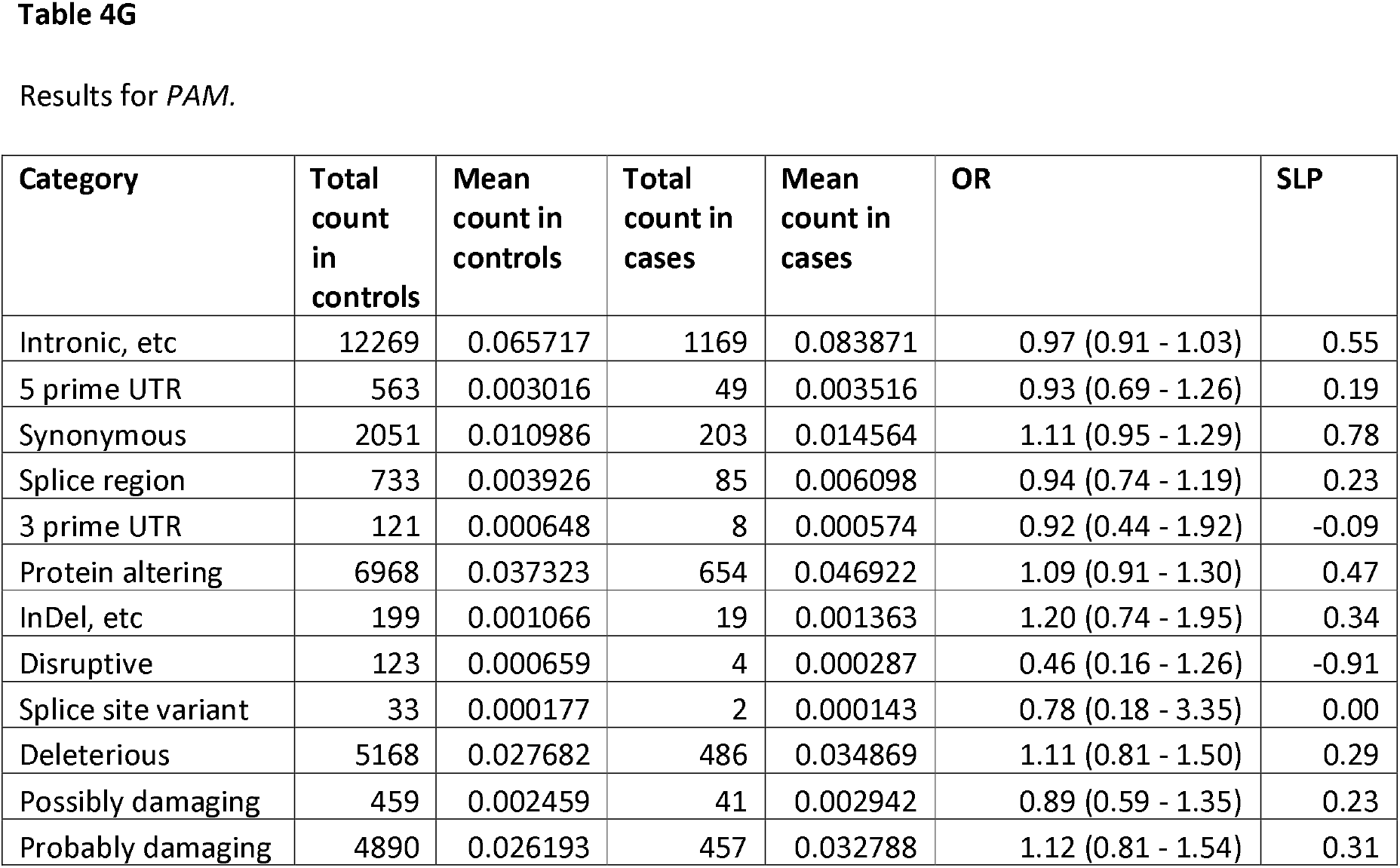
Results from logistic regression analysis showing effect on risk of T2D of different categories of variant within genes previously implicated in diabetes pathogenesis.

| Category | Total count in controls | Mean count in controls | Total count in cases | Mean count in cases | OR | SLP |
| --- | --- | --- | --- | --- | --- | --- |
| Intronic, etc | 8981 | 0.048105 | 798 | 0.057254 | 0.93 (0.86 - 1.00) | 1.24 |
| 5 prime UTR | 413 | 0.002212 | 54 | 0.003874 | 1.26 (0.94 - 1.69) | 0.93 |
| Synonymous | 1561 | 0.008361 | 157 | 0.011264 | 1.05 (0.89 - 1.24) | 0.27 |
| Splice region | 624 | 0.003342 | 87 | 0.006242 | 1.03 (0.81 - 1.31) | 0.08 |
| 3 prime UTR | 330 | 0.001768 | 30 | 0.002152 | 0.78 (0.53 - 1.16) | 0.66 |
| Protein altering | 2806 | 0.015030 | 268 | 0.019228 | 1.07 (0.90 - 1.28) | 0.38 |
| InDel, etc | 8 | 0.000043 | 1 | 0.000072 | 1.96 (0.23 - 16.51) | 0.32 |
| Disruptive | 29 | 0.000155 | 7 | 0.000502 | 3.27 (1.39 - 7.70) | 1.96 |
| Splice site variant | 2 | 0.000011 | 1 | 0.000072 | 7.91 (0.67 - 93.47) | 0.71 |
| Deleterious | 1274 | 0.006824 | 107 | 0.007677 | 0.87 (0.64 - 1.20) | 0.41 |
| Possibly damaging | 430 | 0.002303 | 39 | 0.002798 | 1.07 (0.72 - 1.58) | 0.13 |
| Probably damaging | 449 | 0.002405 | 43 | 0.003085 | 1.27 (0.83 - 1.94) | 0.58 |
| Intronic, etc | 9825 | 0.052626 | 1188 | 0.085235 | 1.03 (0.97 - 1.09) | 0.43 |
| 5 prime UTR | 2883 | 0.015442 | 259 | 0.018582 | 1.08 (0.94 - 1.23) | 0.57 |
| Synonymous | 848 | 0.004542 | 78 | 0.005596 | 1.02 (0.80 - 1.29) | 0.05 |
| Splice region | 140 | 0.000750 | 14 | 0.001004 | 1.44 (0.82 - 2.54) | 0.72 |
| 3 prime UTR | 1116 | 0.005978 | 145 | 0.010403 | 0.89 (0.74 - 1.08) | 0.65 |
| Protein altering | 1734 | 0.009288 | 184 | 0.013201 | 1.19 (0.96 - 1.47) | 1.00 |
| InDel, etc | 7 | 0.000037 | 1 | 0.000072 | 2.04 (0.24 - 17.58) | 0.36 |
| Disruptive | 2 | 0.000011 | 1 | 0.000072 | 6.79 (0.57 - 81.16) | 0.71 |
| Splice site variant | 0 | 0.000000 | 0 | 0.000000 |  | 0.00 |
| Deleterious | 1046 | 0.005603 | 60 | 0.004305 | 0.58 (0.33 - 1.00) | -1.34 |
| Possibly damaging | 591 | 0.003166 | 32 | 0.002296 | 1.00 (0.55 - 1.82) | 0.00 |
| Probably damaging | 445 | 0.002384 | 33 | 0.002368 | 1.39 (0.76 - 2.53) | -0.56 |
| Intronic, etc | 40619 | 0.217570 | 3992 | 0.286411 | 1.02 (0.99 - 1.05) | 0.84 |
| 5 prime UTR | 3981 | 0.021324 | 443 | 0.031784 | 1.06 (0.96 - 1.17) | 0.65 |
| Synonymous | 6894 | 0.036927 | 686 | 0.049218 | 0.98 (0.90 - 1.06) | 0.26 |
| Splice region | 1489 | 0.007976 | 115 | 0.008251 | 0.92 (0.76 - 1.12) | 0.41 |
| 3 prime UTR | 117 | 0.000627 | 6 | 0.000430 | 0.62 (0.27 - 1.45) | -0.58 |
| Protein altering | 5651 | 0.030269 | 521 | 0.037380 | 1.07 (0.93 - 1.23) | 0.49 |
| InDel, etc | 13 | 0.000070 | 1 | 0.000072 | 0.90 (0.11 - 7.17) | 0.00 |
| Disruptive | 103 | 0.000552 | 9 | 0.000646 | 1.19 (0.59 - 2.40) | 0.21 |
| Splice site variant | 58 | 0.000311 | 4 | 0.000287 | 0.83 (0.29 - 2.37) | -0.14 |
| Deleterious | 1861 | 0.009968 | 189 | 0.013560 | 1.17 (0.96 - 1.43) | 0.97 |
| Possibly damaging | 469 | 0.002512 | 38 | 0.002726 | 0.70 (0.48 - 1.02) | 1.26 |
| Probably damaging | 2290 | 0.012266 | 215 | 0.015425 | 1.10 (0.90 - 1.33) | 0.46 |
| Intronic, etc | 36762 | 0.196910 | 3725 | 0.267255 | 0.99 (0.96 - 1.02) | 0.31 |
| 5 prime UTR | 165 | 0.000884 | 15 | 0.001076 | 0.86 (0.50 - 1.49) | 0.23 |
| Synonymous | 15478 | 0.082906 | 1385 | 0.099369 | 1.02 (0.97 - 1.07) | 0.29 |
| Splice region | 4126 | 0.022100 | 387 | 0.027766 | 0.96 (0.82 - 1.13) | 0.22 |
| 3 prime UTR | 400 | 0.002143 | 48 | 0.003444 | 1.24 (0.91 - 1.70) | 0.78 |
| Protein altering | 8249 | 0.044185 | 603 | 0.043263 | 0.89 (0.79 - 1.00) | -1.33 |
| InDel, etc | 18 | 0.000096 | 2 | 0.000143 | 1.41 (0.49 - 4.07) | 0.19 |
| Disruptive | 45 | 0.000241 | 5 | 0.000359 | 1.62 (0.63 - 4.19) | 0.51 |
| Splice site variant | 16 | 0.000086 | 1 | 0.000072 | 0.64 (0.08 - 5.12) | 0.00 |
| Deleterious | 3668 | 0.019647 | 292 | 0.020950 | 1.21 (0.87 - 1.67) | 0.61 |
| Possibly damaging | 3181 | 0.017039 | 249 | 0.017865 | 0.90 (0.65 - 1.25) | 0.27 |
| Probably damaging | 357 | 0.001912 | 26 | 0.001865 | 0.98 (0.59 - 1.62) | -0.03 |
| Intronic, etc | 0 | 0.000000 | 0 | 0.000000 |  | 0.00 |
| 5 prime UTR | 265 | 0.001419 | 25 | 0.001794 | 1.28 (0.84 - 1.96) | 0.63 |
| Synonymous | 799 | 0.004280 | 93 | 0.006672 | 0.97 (0.77 - 1.21) | 0.12 |
| Splice region | 0 | 0.000000 | 0 | 0.000000 |  | 0.00 |
| 3 prime UTR | 45 | 0.000241 | 4 | 0.000287 | 1.26 (0.44 - 3.61) | 0.11 |
| Protein altering | 1248 | 0.006685 | 117 | 0.008394 | 0.91 (0.69 - 1.22) | 0.27 |
| InDel, etc | 3 | 0.000016 | 1 | 0.000072 | 4.85 (0.48 - 49.39) | 0.60 |
| Disruptive | 72 | 0.000386 | 8 | 0.000574 | 1.47 (0.69 - 3.12) | 0.51 |
| Splice site variant | 0 | 0.000000 | 0 | 0.000000 |  | 0.00 |
| Deleterious | 412 | 0.002207 | 44 | 0.003157 | 1.22 (0.61 - 2.44) | 0.25 |
| Possibly damaging | 181 | 0.000970 | 22 | 0.001578 | 1.39 (0.78 - 2.49) | 0.59 |
| Probably damaging | 389 | 0.002084 | 39 | 0.002798 | 1.28 (0.60 - 2.72) | 0.29 |
| Intronic, etc | 7313 | 0.039171 | 607 | 0.043550 | 0.93 (0.85 - 1.02) | 0.88 |
| 5 prime UTR | 307 | 0.001644 | 36 | 0.002583 | 1.21 (0.85 - 1.74) | 0.55 |
| Synonymous | 509 | 0.002726 | 65 | 0.004664 | 1.40 (1.07 - 1.84) | 1.92 |
| Splice region | 351 | 0.001880 | 40 | 0.002870 | 1.12 (0.80 - 1.58) | 0.31 |
| 3 prime UTR | 2262 | 0.012116 | 197 | 0.014134 | 1.04 (0.88 - 1.23) | 0.18 |
| Protein altering | 2666 | 0.014280 | 257 | 0.018439 | 0.96 (0.80 - 1.14) | 0.19 |
| InDel, etc | 3 | 0.000016 | 0 | 0.000000 |  | 0.00 |
| Disruptive | 99 | 0.000530 | 9 | 0.000646 | 0.82 (0.40 - 1.68) | 0.23 |
| Splice site variant | 59 | 0.000316 | 1 | 0.000072 | 0.24 (0.03 - 1.77) | -0.82 |
| Deleterious | 616 | 0.003300 | 30 | 0.002152 | 0.61 (0.29 - 1.30) | -0.72 |
| Possibly damaging | 455 | 0.002437 | 26 | 0.001865 | 0.89 (0.57 - 1.39) | -0.23 |
| Probably damaging | 441 | 0.002362 | 22 | 0.001578 | 0.98 (0.41 - 2.34) | -0.02 |
| Intronic, etc | 12269 | 0.065717 | 1169 | 0.083871 | 0.97 (0.91 - 1.03) | 0.55 |
| 5 prime UTR | 563 | 0.003016 | 49 | 0.003516 | 0.93 (0.69 - 1.26) | 0.19 |
| Synonymous | 2051 | 0.010986 | 203 | 0.014564 | 1.11 (0.95 - 1.29) | 0.78 |
| Splice region | 733 | 0.003926 | 85 | 0.006098 | 0.94 (0.74 - 1.19) | 0.23 |
| 3 prime UTR | 121 | 0.000648 | 8 | 0.000574 | 0.92 (0.44 - 1.92) | -0.09 |
| Protein altering | 6968 | 0.037323 | 654 | 0.046922 | 1.09 (0.91 - 1.30) | 0.47 |
| InDel, etc | 199 | 0.001066 | 19 | 0.001363 | 1.20 (0.74 - 1.95) | 0.34 |
| Disruptive | 123 | 0.000659 | 4 | 0.000287 | 0.46 (0.16 - 1.26) | -0.91 |
| Splice site variant | 33 | 0.000177 | 2 | 0.000143 | 0.78 (0.18 - 3.35) | 0.00 |
| Deleterious | 5168 | 0.027682 | 486 | 0.034869 | 1.11 (0.81 - 1.50) | 0.29 |
| Possibly damaging | 459 | 0.002459 | 41 | 0.002942 | 0.89 (0.59 - 1.35) | 0.23 |
| Probably damaging | 4890 | 0.026193 | 457 | 0.032788 | 1.12 (0.81 - 1.54) | 0.31 |

## Discussion

These analyses provide a broad overview of the way very rare genetic variants contribute to risk of type 2 diabetes in a large sample broadly representative of the population. The weighted burden analysis successfully identifies two known diabetes genes, *GCK* and *HNF4A*, and implicates a novel gene, *GIGYF1*. A few other biologically plausible genes do not reach formal levels of statistical significance after correction for multiple testing but might be worthy of further investigation, including *ALAD, PPARG, GYG1, GHRL, SCL2A2* and *SLC2A4*. Any possible role for these genes will become clearer as additional data becomes available, for example from the remaining 300,000 UK Biobank subjects for whom exome sequence data has not yet been released. Typically, hundreds of variants are identified per gene, mostly occurring in only a handful of subjects each. The findings for some previously implicated genes, *HNF1A, HNF1B, ABCC8, MC4R* and *SLC30A8*, are not formally statistically significant but are consistent with an effect which could be detected once the sample size is increased.

Together, variants which can be identified as having large effects on risk occur in fewer than 1% of the cases of T2D in this sample. There is no doubt that identifying specific genetic causes may be useful to guide treatment for some patients (Agostini et al., 2018). However it needs to be acknowledged that for the vast majority of patients with T2D exome sequencing will be not be helpful in terms of identifying specific subtypes of disease which might benefit from specific treatments. Thus, the potential to apply a personalised medicine approach to T2D based on genetic testing seems to be somewhat limited.

The main potential utility of genetic investigations such as this might be to better characterise the mechanisms which can lead to disease, identify novel drug targets and develop improved therapeutic approaches which would benefit T2D patients in general, rather than only the small number carrying the relevant genetic variant. If some of the tentative findings reported here can be replicated then the genes identified could become the objects of more intensive investigation.

## Data availability

The raw data is available on application to UK Biobank. Detailed results with variant counts cannot be made available because they might be used for subject identification. Scripts and relevant derived variables will be deposited in UK Biobank.

## Acknowledgments

This research has been conducted using the UK Biobank Resource. The author wishes to acknowledge the staff supporting the High Performance Computing Cluster, Computer Science Department, University College London. This work was carried out in part using resources provided by BBSRC equipment grant BB/R01356X/1. The author wishes to thank the participants who volunteered for the UK Biobank project.

